# ADHD symptom reduction after following an FFD is associated with gut microbiome composition

**DOI:** 10.1101/2024.07.11.24310255

**Authors:** S Hontelez, M Guthrie, T Stobernack, P van Baarlen, C Rousseau, MP Boks, R Rodrigues Pereira, J Boekhorst, M Kleerebezem

**Affiliations:** Host-Microbe Interactomics, Wageningen University and Research, De Elst 1, 6708 WD, Wageningen, The Netherlands.; Department of Psychiatry, Brain Center University Medical Center Utrecht, University Utrecht, Utrecht, The Netherlands; Medical Centre Kinderplein, Rotterdam, The Netherlands

## Abstract

Attention-deficit hyperactivity disorder (ADHD) is one of the most common childhood neuropsychiatric conditions. Both (epi)genetic and environmental factors are suggested to contribute to the aetiology of ADHD. In the last decade, nutrition has received considerable attention as potential environmental factor triggering ADHD behaviour. Studies applying a few-foods diet (FFD) can lead to behavioural improvements of at least 40% in 50-64% of children with ADHD. It is conceivable that the Microbiota-Gut-Brain (MGB) axis is involved in the mechanism of action underlying the behavioural improvements observed in children with ADHD after following the FFD. This study investigated potential associations between changes in ADHD symptoms in children that followed an FFD. We identified a significant association between microbiota composition and change in ADHD symptoms in food-associated ADHD.

## Introduction

Attention-deficit hyperactivity disorder (ADHD) is one of the most common childhood neuropsychiatric conditions, with 6% prevalence world-wide^1^. The incidence is increasing rapidly; in the U.S., the prevalence of ADHD in children and adolescents has increased from 6.1 in 1997 to 10.2 percent in 2016^2^. In Europe including the Netherlands, ranges are 3 to 4-5%^3^ ADHD is typically characterised by a combination of inattentive, hyperactive and impulsive behaviour^4^ and is an impairing condition that is burdensome for child, family and society. Notably, ADHD behaviour symptoms are highly variable with considerable variations in the relative degree of inattentive and hyperactive behaviours^3^.

Although the cause(s) underlying ADHD is (are) still unclear, it is commonly accepted that both (epi)genetic and environmental factors contribute to the aetiology of ADHD^5^. An environmental factor that has received considerable attention in the last decade is nutrition. Clinical trials studying the effect of food ingredients and/or additives on ADHD symptom severity have reported small and often inconsistent or clinically irrelevant effects on behaviour. Conversely, studies applying a diet that allows only a few foods, the so-called few-foods diet (FFD), resulted in a clinically relevant reduction of ADHD symptoms in children following the FFD^6^, resulting in behavioural improvements of at least 40% in 50-64% of children with ADHD^7–9^. Considering the evidence for the effect of an FFD on ADHD symptoms, we initiated the ‘Biomarker Research in ADHD: the Impact of Nutrition’ (BRAIN) study with the aim to investigate the mechanisms underlying the behavioural improvements after following an FFD. In this open-label trial, 63% of the participants responded with behavioural improvements of at least 40% after following the FFD, with an average improvement of 73%. The decrease in parent-reported ADHD symptoms was correlated with an activity increase in the precuneus region of the brain during performance of a response-inhibition task^10^.

Since brain function can be affected by the microbiome ^11^, which in turn is strongly affected by diet^12,13^, it is conceivable that the Microbiota-Gut-Brain (MGB) axis is involved in the mechanism of action underlying the behavioural improvements observed in children with ADHD after following the FFD. For example, specific microbial metabolites that can be transported via the vagus nervus to brain might influence (severity of) ADHD^14^. Moreover, the well-established personalized composition of the gut microbiome (REF) may partially explain the large variation observed in behavioural responses in children with ADHD when subjected to an FFD intervention.

This study aimed to investigate potential associations between changes in ADHD symptoms and MGB parameters (i.e., gut microbiota composition and functionality, metabolome, blood immune cell transcriptome and [epi]genetic background) after following an FFD in children who participated in the BRAIN study. While the type of intervention, i.e., a diet strictly limited to a small group of food ingredients, does not allow for a straightforward double-blind placebo-controlled study, the longitudinal setup and direct molecular readouts allow quantification of the role of the gut microbiota composition and genetic content and allowed us to assess if specific host parameters act as mediators between diet and ADHD symptoms.

## Results

Of the 100 BRAIN study participants, 21 were excluded due to non-compliance, or stopped prematurely with the diet (figure 1). Inclusion / exclusion criteria are reported in the methods section. During progressive reduction of allowed food ingredients (progressing from FFD-E1 to FFD-E2 to FFD; see methods for details) in a period of 5 weeks, six children responded favourably to the FFD.E1 and five children responded to the FFD.E2 (figure 2). For those children that showed a favourable response, the diet was not further restricted. For 68 children the FFD.E1 and FFD.E2 were not effective, therefore these children proceeded with the FFD. For all 79 children, faecal, urine and buccal swap samples were collected before (t1) and after (t2) the diet intervention (table 1). Blood sample sets (t1+t2) for metabolite profiling and peripheral blood mononuclear cells (PBMC) isolation were complete for 76 and 67 children, respectively. We obtained eight omics datasets from faecal, urine, buccal swab samples and blood samples. Gut microbiome composition and functionality were derived from analysis of next-generation sequencing results (shotgun sequencing and 16S profiling) for DNA extracted from the faecal samples. Relative metabolite levels were measured in plasma and urine samples. Gene expression profiles were obtained from blood derived PBMCs. SNPs and methylated DNA loci profiles were derived from the buccal swaps (table 1). ADHD rating scale (ARS) was assessed during the screening (t0), at t1 and at t2, showing consistent scores between t0 and t110 while ARS scored reduced considerably from t1 to t2 (Figure 2b). Of the 79 children, 50 (63%) showed a reduction in ARS score of ≥40% and were therefore categorised as responders.

**Figure 1:**
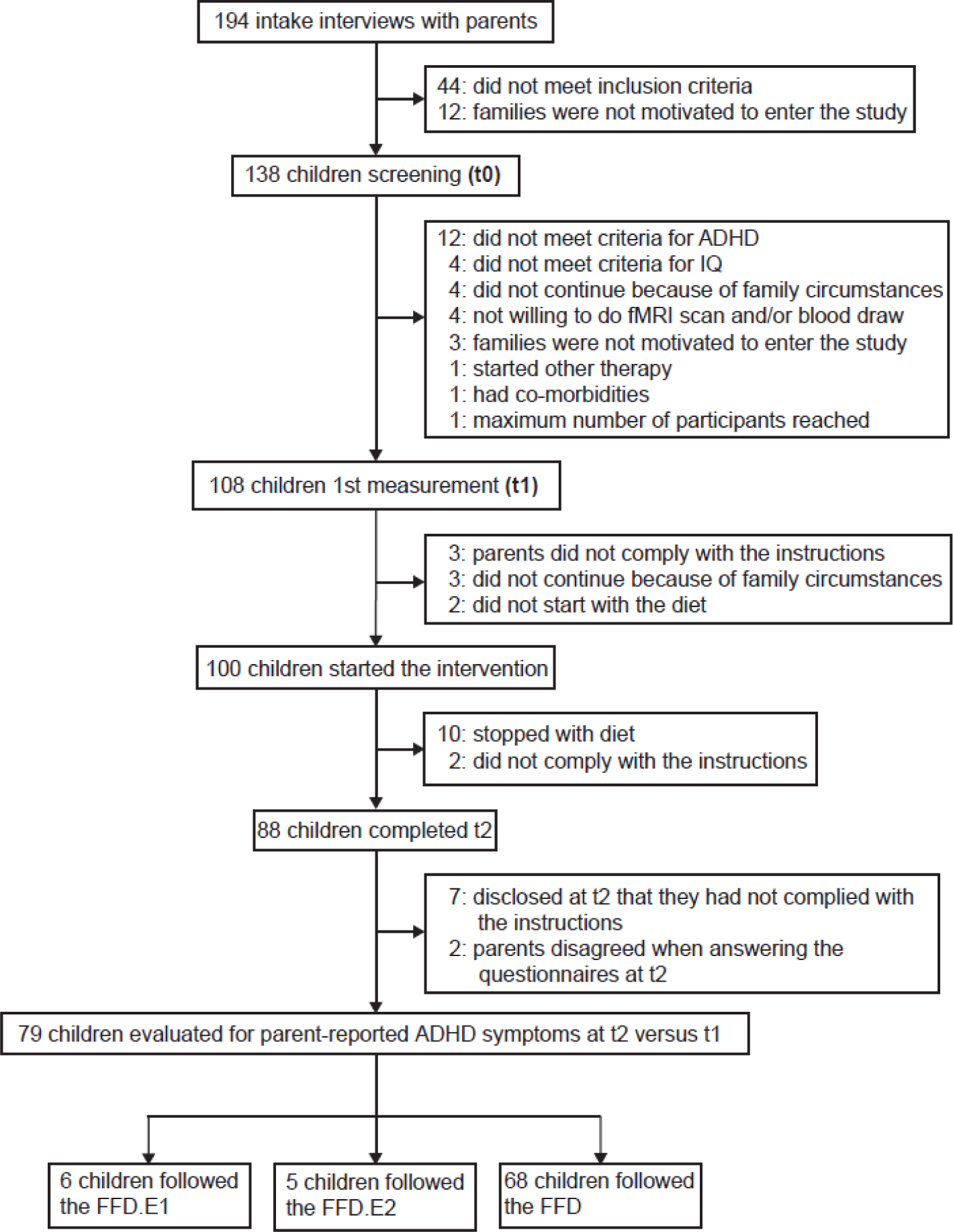
Flowchart BRAIN study participants.

**Figure 2:**
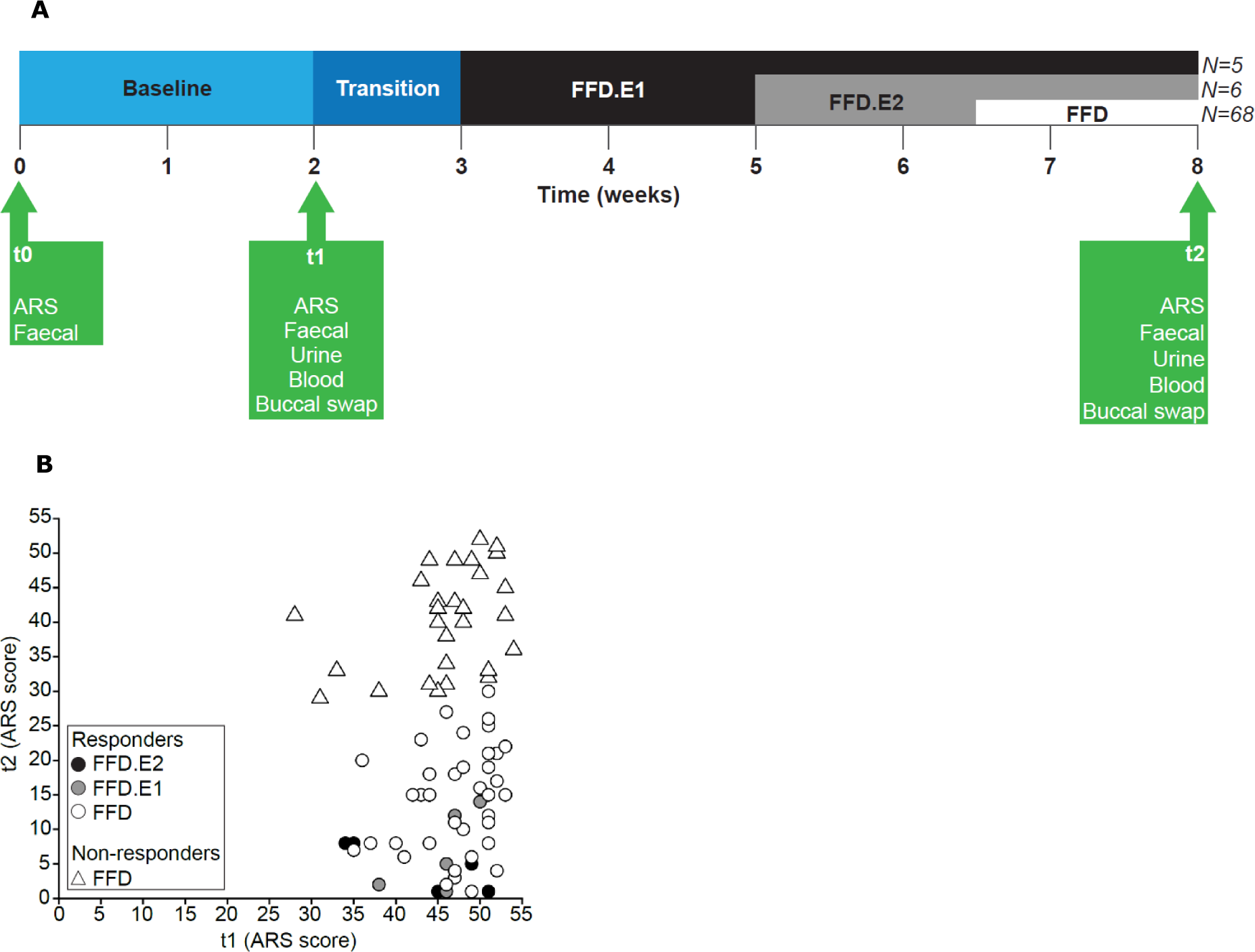
ADHD symptom reduction after diet intervention. A) Study design. B) ARS score at t1 vs t2 of all children included for analysis (n=79).

**Table 1.** Overview of collected samples and data sets.

| View | Source | Samples (n) |  |  | Features (n) |  |
| --- | --- | --- | --- | --- | --- | --- |
|  |  | t1 | t2 | t1 & t2 | Total | MOFA |
| 16S (V3-V4; genera) | Faeces | 79 | 79 | 79 | 210 | 210 |
| Metagenomics species | Faeces | 79 | 79 | 79 | 332 | 332 |
| Metagenomics genefamilies | Faeces | 79 | 79 | 79 | 2967555 | 4999 |
| Plasma metabolites (named/unnamed) | Blood | 77 | 76 | 76 | 852 / 215 | 852 / 215 |
| Urine metabolites (named/unnamed) | Urine | 79 | 79 | 79 | 904 / 421 | 904 / 421 |
| Gene expression PBMCs (genes)<sup>1</sup> | Blood | 74 | 71 | 67 | 19176 | 5003 |
| SNPs | Buccal swaps | 79 | 79 | 79 | 654027 | 4785 |
| DNA methylation (loci) | Buccal swaps | 71 | 77 | 70 | 850000 | 5000 |

<sup>1</sup> With at least 1 mapped read in 1 sample.

### Phenylalanine and tyrosine metabolism

In the published study protocol^15^, we proposed that after adhering to FFD, there might be correlation between ARS score and levels of specific metabolites that had been previously associated with ADHD. We assessed potential associations between ARS change (categorical and continuous) and the subset of data defined as primary outcome measures in the study protocol: phenylalanine and tyrosine levels in blood and urine, and abundance of 21 gut microbial genes involved in phenylalanine and tyrosine metabolism. Spearman rank analyses (continuous) and Mann-Whitney tests (categorical) of the gene relative abundances measured with feacal metagenomics sequencing and metabolite levels with ARS change showed no significant correlations (table S1, S2).

### Diet type affects metabolites and microbiota composition

To find cross-omics variation associated with relative change in ADHD symptoms (ARS change) between pre- and post-FFD timepoints, we employed the Multi Omics Factor Analysis (MOFA) method. MOFA can be viewed as a versatile and statistically rigorous generalization of principal component analysis (PCA) to multi-omics data, separating the omics features into factors according to common variation^16^. Eight data sets (referred to as “views” in the context of MOFA) were used, with a maximum of ∼5000 features per view (table 1).

We assessed whether any of the 10 factors in the MOFA model associated with ARS change, timepoint (t1 vs t2) or type of diet (FFD.E1, FFD.E2 or FFD; figure 3a). Both the timepoint (i.e., the pre- and post-intervention measurements) and the diet type were significantly associated with factor 2, while none of the factors correlated with ARS change. Microbiome and metabolite features provided the largest contribution to the factor values in factor 2 (figure 3b). The difference between t1 and t2 factor 2 values indicated that the features of this factor had changed considerably between t1 and t2 (figure 3c). The effect was largest for the FFD and smallest for the FFD.E1, suggesting that the foods that were allowed in the diet affected the metabolites and microbiome features in this factor. Several metabolites with higher levels at t2 compared to t1 were related to consumption of poultry (1-methyl-5-imidazoleacetate, anserine, (N-acetyl-)3-methylhistidine; figure 4a), concurrent with the FFD, during which only turkey meat is allowed (FFD.E1/E2 also allows lamb). Metabolites that were detected at t1 but not at t2 are often derived from foods not allowed during the FFD (i.e., theobromine [chocolate], thymol sulphate [herbs], 3/7-methylxanthine [caffeinated foods/drinks], piperine [pepper]). Comparing the three diet types by RDA further shows that cereal-derived metabolites (2-aminophenol and 2-acetamidophenol) are enriched in children that followed the FFD.E1 diet type that allowed consumption of wheat-based products (figure 4b,c); these products were not allowed in the FFD.E2 and FFD diet types. Piperine metabolites were detected in both FFD.E1 and FFD.E2 children. Pepper was allowed in both these diet types but excluded for children following the FFD. The homogeneous change in factor values of factor 2 coinciding with the change in diet, as well as the discriminating metabolites related to dietary intake, are further corroborating that compliance to the dietary instructions by the participants was high.

**Figure 3:**
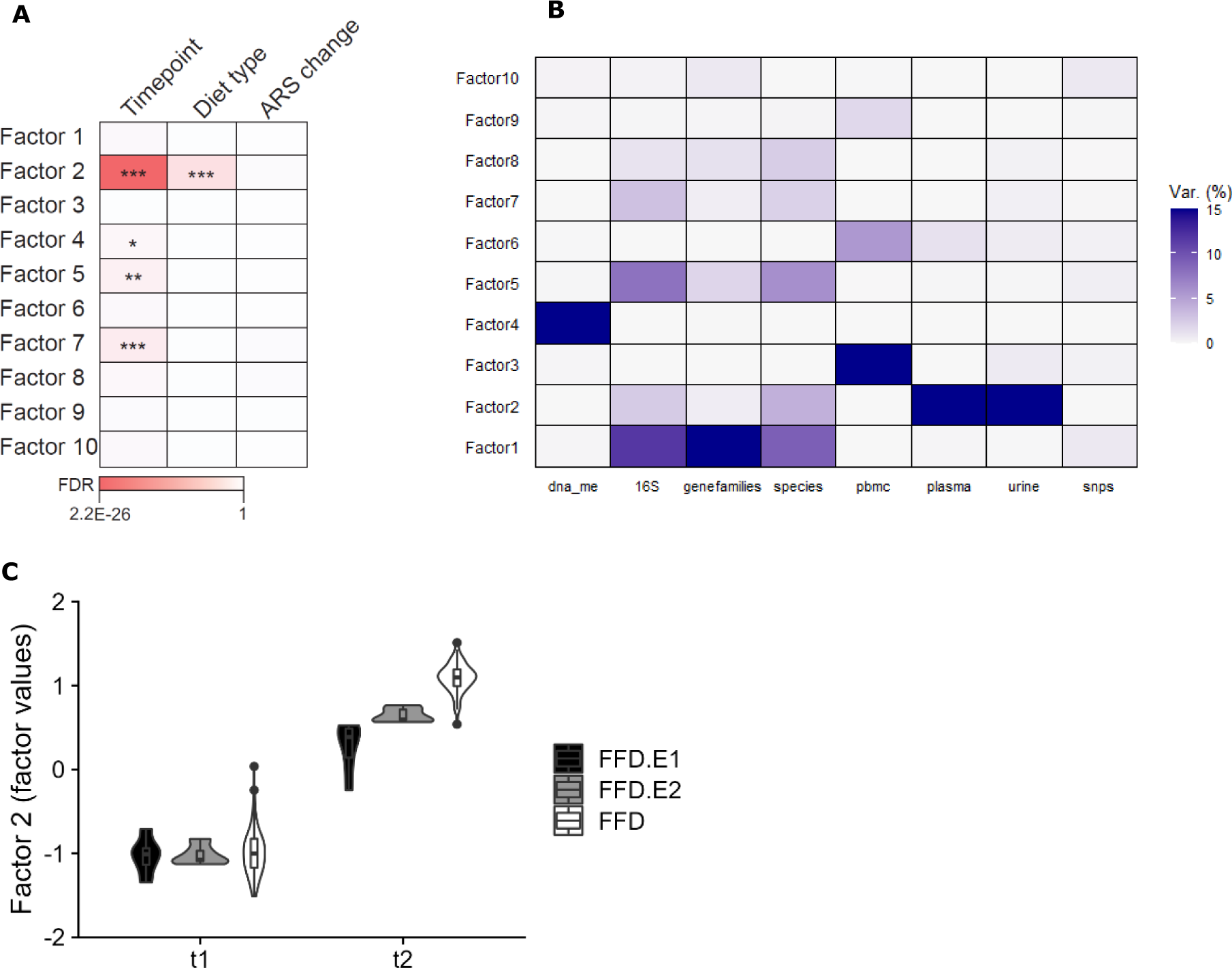
10-factor MOFA model (n=79). A) Associations of the 10 factors in the MOFA model with timepoint (t1 vs t2), diet type (FFD.E1, FFD.E2 or FFD) and ARS change (%). B) Explained variation (%) of the data views per factor. C) Factor 2 values changes between t1 and t2 for the 3 diet types

**Figure 4.**
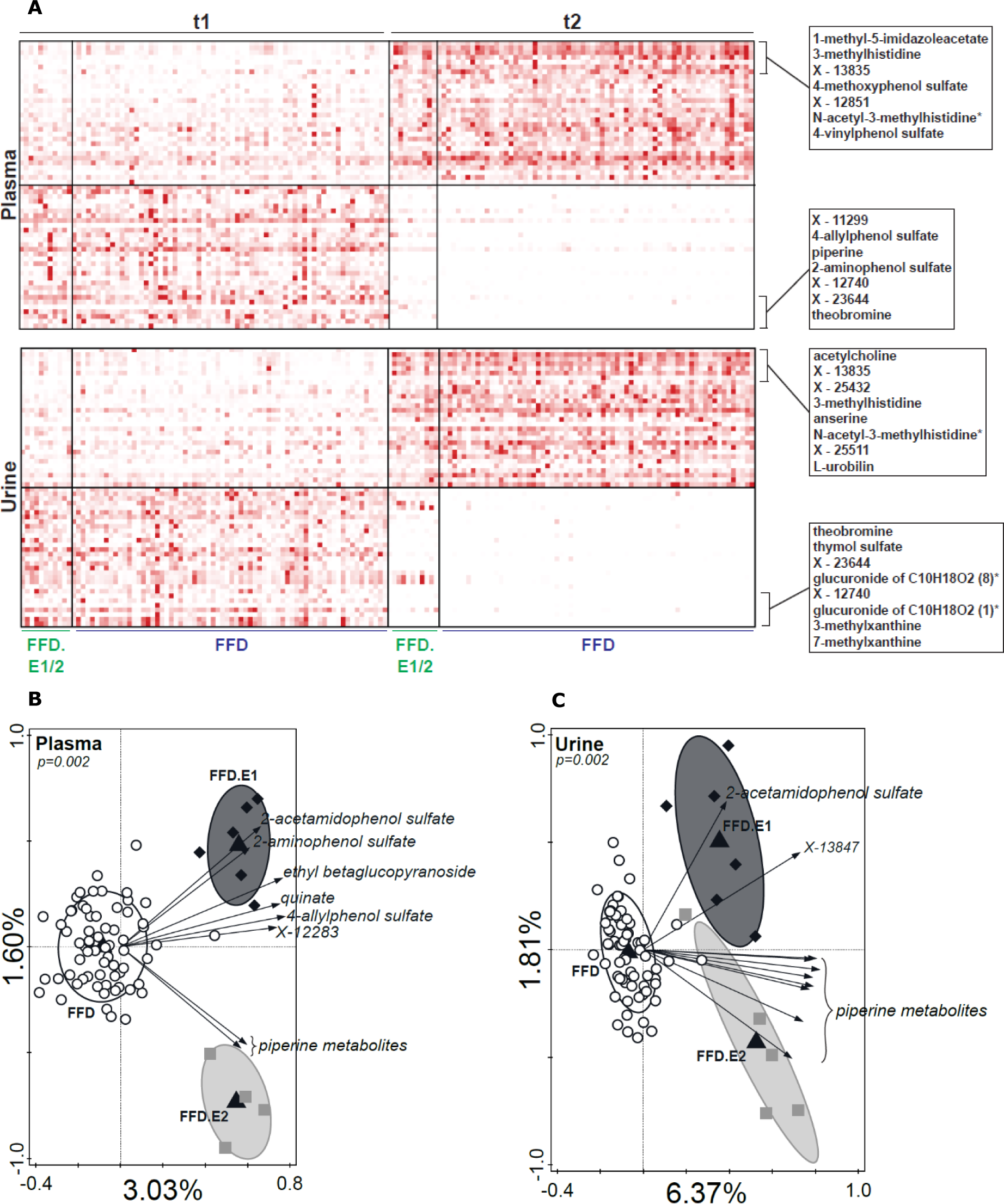
Metabolites that decreased or increased in abundance after following the FFD. A) Heatmap of metabolite levels in blood and urine that showed the largest contribution to MOFA factor 2. B,C) RDA of plasma (B) and urine (C) metabolites with diet type as explanatory variable at t2.

Because children that responded favourably to the FFD.E1/E2 diets all showed a high ARS change (figure 2b) and had a smaller shift in factor 2 values from t1 to t2 compared to the FFD diet types (figure 3c), diet type could have confounded the analyses between the MOFA factors and ARS change. Therefore, we generated MOFA models using only the data of the children that followed the FFD. Like in the models with all children, factor 2 captured the t1-t2 change in metabolites, however, none of the MOFA factors, including factor 2, significantly associated with ARS.

### ARS change correlated with gut microbiome composition

Potential associations between ARS change and the variation in the omics data views were assessed using a supervised RDA approach. RDA analyses were performed using only the data derived from the children that followed the FFD diet type; children following the FFD.E1 and FFD.E2 were added supplementary in the RDA plot. Both t1 and t2 timepoints were included, permuting per participant. Species abundance was significantly associated with ARS change (p=0.008, explained variation=2.07%, figure 5a, table 2). The supplementary FFD.E1 and FFD.E2 samples appeared scattered across the RDA space, while, on basis of the high ARS change of with these samples, they would have been expected to be positioned towards the far-right hand side of the plot. Therefore, the observed association between species abundance and ARS holds for the children that responded to the FFD, but not for the children that responded already to FFD-E1 or FFD-E2, justifying the separate investigation of the FFD group. Within the latter group, species such as *Bacteroides dorei*, *Akkermansia municiphila*, and *Alistipes onderdonkii* were more abundant in children with a low ARS change, while *Dorea formicigenerans*, *Roseburia inulinivorans* and *Ruminococcus torques* were more abundant in children with a high ARS. Performing RDAs per timepoint confirms that the contribution (horizontal direction arrows) in separate RDAs for t1 and t2 was very similar (figure S2).

**Figure 5.**
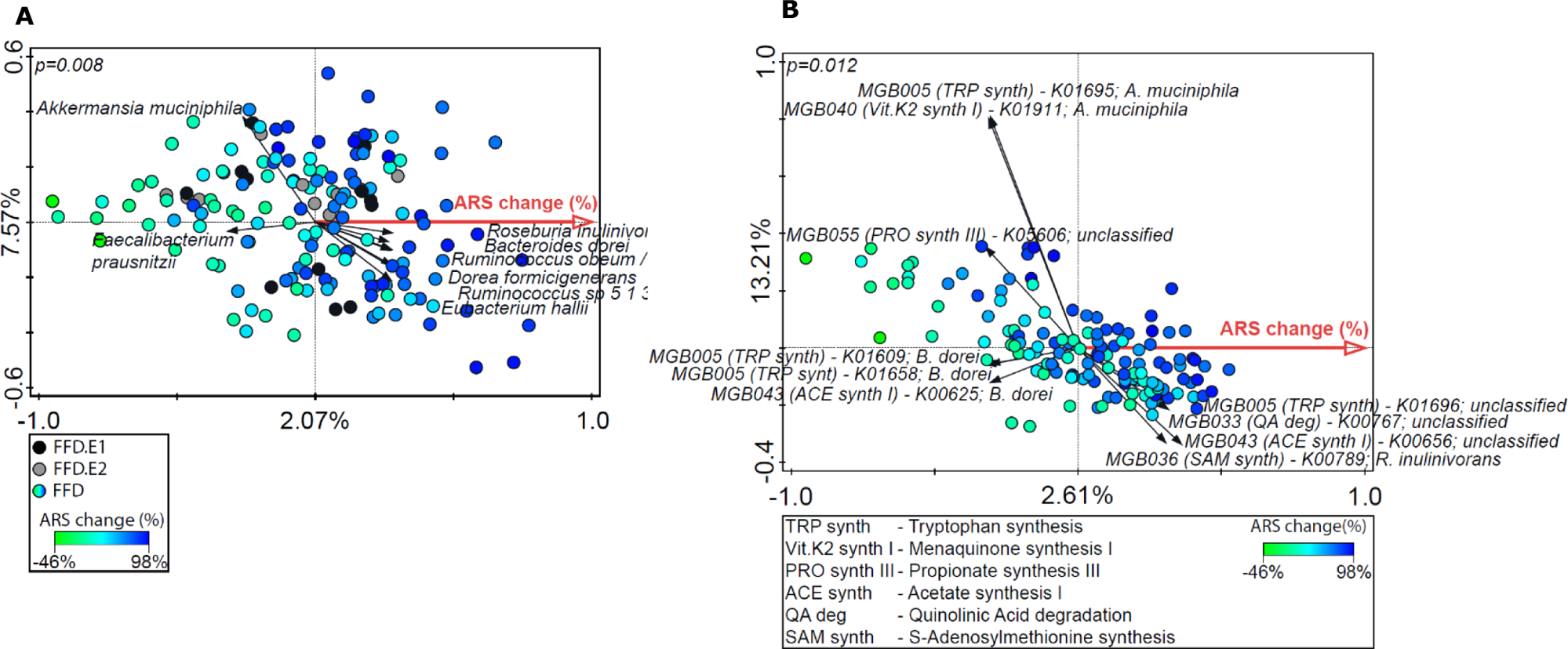
Association between microbiome and ARS change in children that followed the FFD (n=68). RDAs with ARS change as explanatory variable and A) relative gut microbiome species abundance at t1 and t2 or B) relative abundance of MGB KOs stratified to gut microbiome species as response variable. In both plots the axis labels depict the percentage explained variation of the RDA-axis (x-axis) and the first PCA axis (y-axis). Permutation was performed between subjects rather than between samples.

**Table 2.** RDA results with ARS change as explanatory variable and the omics data views as response variables.

| Data view | Timepoint | N samples | Explained variation (%) | p-value<sup>1</sup> | FDR<sup>2</sup> |
| --- | --- | --- | --- | --- | --- |
| 16S (V3-V4) | t1+t2 | 136 | 1.94 | 0.012 | 0.048 |
| Metagenomics Species abundance | t1+t2 | 136 | 2.07 | 0.008 | 0.048 |
| Metagenomics Genefamilies | t1+t2 | 136 | 0.72 | 0.572 | 0.654 |
| Plasma metabolites | t1+t2 | 132 | 1.01 | 0.45 | 0.653 |
| Urine metabolites | t1+t2 | 136 | 0.87 | 0.49 | 0.653 |
| SNPs | t1 | 68 | 1.42 | 0.358 | 0.653 |
| DNA methylation | t1+t2 | 126 | 1.42 | 0.104 | 0.277 |
| PBMC RNA expression | t1+t2 | 126 | 0.72 | 0.662 | 0.662 |

FDR=false discovery rate; SNP=single nucleotide polymorphism; PBMCs=peripheral blood mononuclear cells
<sup>1</sup> Monte Carlo permutation test, Canoco5
<sup>2</sup> Benjamini & Hochberg

To predict functional impact of the microbial species identified, we employed the gut-brain modules that represent gut-microbial pathways with potential neuroactive effects ^17^. Abundance of the genes that make up the modules, stratified to species was used to test whether the species associated with ARS change represented specific modules. RDA with the stratified genes as response variable and ARS change as explanatory variable found a significant association (p=0.012, explained variation=2.61%, figure 5b). The explained variation was 0.54% higher than the RDA with species abundance as response variable, suggesting that functionality adds information that is relevant to the variation related to ARS change.

The contribution (RDA-axis, i.e., horizontal direction arrows) of the stratified genes to the RDA ordination shows that most genes have both positive and negative coordinates (i.e., these stratified genes were enriched in microbiota samples from children with high and low ARS change, respectively) depending on the species by which they are encoded (figure S3a). Likewise, comparing the RDA-axis contribution of the microbial species between the species RDA and stratified Kegg Orthology (KO) RDA shows that specific species can be enriched both in high and low responders, depending on the genes that they encode (figure S3b).

### Peripheral changes related to ARS change

Subsequently we investigated whether the change in ARS score (figure 2b) was associated with changes in blood-metabolite levels and PBMC transcription profiles, using EdgeR and Ingenuity Pathway analysis (IPA). These analyses revealed only modest correlations with differential (increased) gene expression and metabolite concentrations involved in among others amino acid metabolism mitochondrial respiration, suggesting that the microbiome association with ARS score changes are not clearly reflected in the blood samples collected.

### Association between microbiome composition, behaviour and brain activation

Finally, we assessed the relation between behaviour, microbiome composition and brain activation. In our previous publication^10^, we showed that ARS change was positively correlated with activation of the precuneus during execution of a stop-signal task, applying two response inhibition contrasts (StopSuccess>Go: general linear model [GLM], df=50, n=53, pFWE=0.015; StopSuccess>StopFail: GLM, df=50, n=53, pFWE<0.001). To investigate whether the ARS change and correlated precuneus activation was also associated with microbiome species composition, the cluster-averaged beta weights for both response-inhibition contrasts were used as explanatory variables in RDAs with relative gut microbiome species composition as response variable (figure 6a, b). RDA found that Beta weights of precuneus activation in both contrasts were significantly associated with species relative abundance.

**Figure 6.**
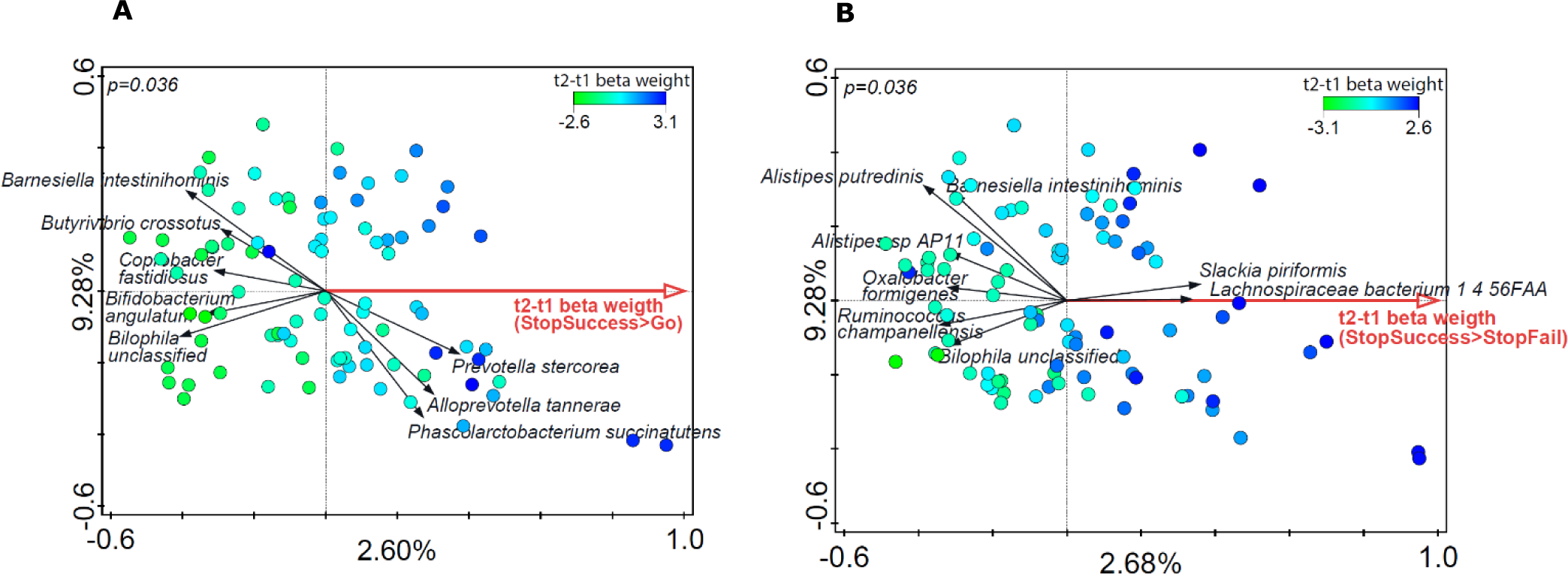
Association between precuneus activation after FFD and microbiome species abundance (n=45). A-B) RDAs with microbiome species abundance (both timepoints) as response variable and precuneus t2-t1 beta weights of A) the StopSuccess>Go or B) the StopSuccess>StopFail contrasts. The axis labels depict the percentage explained variation of the RDA-axis (x-axis) and the first PCA axis (y-axis). Permutation was performed between subjects rather than between samples.

## Discussion

This study aimed to unravel the MGB mechanisms potentially underlying reduction in ADHD symptoms in children with ADHD after following an FFD. Metabolite levels of tyrosine and phenylalanine, and relative abundances of the 21 microbial enzymes involved in their metabolism were not associated with changes in ADHD symptoms after following the diet. However, we found that composition of the gut microbiome was associated with change in ADHD symptoms as well as our previously published activation of the precuneus brain region.

Exploratory analysis of the data revealed a substantial impact of the diet on specific metabolites and relative abundance of specific gut microbiota taxa. Notably, the three different FFD types each presented a signature effect on blood and urine metabolites, with changes in specific metabolites that could be related to the foods allowed or excluded in the respective FFD types. The change in levels of these discriminating metabolites in children after following the three diet types suggest that compliance to the dietary instructions by the participants was high. To exclude a confounding effect of the diet type followed by the children, subsequent analyses only included the 68 children that followed the FFD (excluding the 11 children on the FFD.E1/2). While the unsupervised Multi-Omics Factor Analysis (MOFA) approach did not find associations between change in ADHD symptoms and the eight data sets derived from 8 different omics types, applying the supervised RDA method on the individual data sets showed an association between ADHD symptom change and gut microbiome species composition. This may be interpreted to suggest that the variation in the omics data that is associated with behaviour change had been too small to be detected using unsupervised methods, presumably because of genetic and molecular variation between individual children. Children that followed the FFD.E1/2 did not comply with this association, justifying separate statistical investigation by RDA of only the children that followed the FFD. Interestingly, the ARS change associated species composition was very similar before and after the following the FFD. These results could suggest that the gut microbiome plays a role in food-associated ADHD, and that replacing ADHD-associated diets with an FFD had taken away dietary compounds that those bacterial proteins had acted upon or, had introduced FFD-associated compounds that improved ADHD symptoms via a microbiome-gut-brain axis.

Despite the apparent association between microbiome, brain and behaviour, our gene expression data derived from circulating peripheral blood monocytes and blood metabolome data did not point towards peripheral factors in blood that might mediate these associations in a gut-brain axis. It cannot be excluded that peripheral blood transports locally present metabolites or chemical messengers from the intestine to the brain, but that our timing of blood sampling did not allow us to sample such messengers (nor any associated gene expression changes). For example, food-associated metabolites that trigger ADHD symptoms may have been cleared from blood after overnight fasting, such that any food-associated metabolites were not sampled in the morning at sampling time. Also, urine metabolites collected at the same timepoint failed to propose physiological changes associated with behaviour changes. An alternative explanation of the behaviour changes because of the FFD diet could involve the enteric nervous system (ENS) that could react to local, mucosal changes in metabolites or gene expression that affect ENS activity in a way that influences brain activity (e.g., precuneus activation) and ADHD symptoms.

We expect that human individuality, i.e., genetic and molecular physiological differences (including microbiome) between the children have complicated identification of factors consistently changing in concert with the behavioural outcome measures. For example, changes in microbiome because of the FFD depend in part on the composition of the microbiome at baseline. Notwithstanding our selection criteria, standing genetic and microbiome differences between the children could have led to loss of power to identify significant correlations between separate omics datasets and changes in ADHD symptoms after following FFDs. MOFA was designed to ameliorate this problem by looking for combinations of features (“factors”) from different omics datasets that jointly capture relatively large fractions of the total variation in the data (“principal component analysis across datasets”). While MOFA did identify combinations of factors from multiple datasets, these combinations reflected either the impact of the diet found in blood and urine metabolomes (figure 3B, factor 2), or correlations between microbiota composition and microbiota gene families (e.g., figure 3B, factor 1). No factors with a significant correlation to ADHD symptom changes were found by MOFA.

The strength of the present study is that it confirms earlier reported beneficial effects of FFD diets on ADHD-behavioural symptoms in children, supporting that at least part of childhood ADHD could be diet induced and as such remedied by diet adjustment. In addition, the study collected a range of lowly-invasive samples of the participating children to assess whether broad-spectrum omics analyses could reveal a mechanism underlying such diet-induced ADHD. The pre-diet microbiome composition is linked to the responsiveness of the participants to the FFD diet, and this signature is observed at both t1 and t2. Moreover, extensive *in silico* investigation of the metagenome revealed that the species-specific distribution of specific enzymes and pathways that were previously categorized as potential microbiome gut-brain modules^17^ showed statistical correlations with the measured ARS changes reflecting behaviour. Our omics data sets can be used to underpin diet-induced changes in (ADHD) behaviour with molecular data reporting on changes in microbiome composition and corresponding changes in host circulating blood metabolites, gene expression of peripheral blood immune cells and their methylation status. This wealth of data from children and their microbiome, obtained by different unbiased omics techniques, comes with metadata that allow further in silico analysis once novel statistical analytical techniques and annotations become available.

## Methods

### Participants

This study includes 79 participants of the Biomarker Research in ADHD: the Impact of Nutrition (BRAIN) study^10,15^. Right-handed boys, aged ≥8 and ≤10 years, and meeting the Diagnostic and Statistical Manual of Mental Disorders, fourth edition (DSM-IV) criteria of ADHD were recruited via the media and healthcare institutions. Exclusion criteria were (i) diagnosis of autism spectrum disorder, developmental coordination disorder, chronic gastrointestinal disorder, autoimmune disorder, dyslexia or dyscalculia, (ii) premature birth (<36 weeks) and/or known oxygen deprivation during birth, (iii) vegetarian/vegan, (iv) IQ<85, (v) use of systemic antibiotics, antifungals, antivirals or antiparasitics in the past six months, (vi) insufficient command of the Dutch language by either parents or child, (vii) family circumstances that may compromise compliance, and (viii) having a contraindication to MRI scanning. Participants were allowed to withdraw from the study at any time. All parents gave written informed consent, and all children gave written assent. The required samples size for the primary hypothesis (i.e., association between ARS change and relative abundance of 21 gut microbial ECs and tyrosine and phenylalanine levels in blood plasma and urine) of this manuscript was estimated at 46^15^.

### Study design

The study design of the BRAIN study has been described in detail in previous publications^10,15^. In brief, after the intake eligible children were invited for the screening session (t0) at Wageningen University and Research, the Netherlands. Children meeting the inclusion criteria started with a two-week baseline period adhering to their regular diet, while parents documented food intake and behaviour. After the baseline period (i.e., at t1), blood, urine, stool and buccal swap samples were collected, and ADHD symptom scores were assessed. A one-week transition period followed after t1, gradually adapting the diet to habituate to a different eating pattern. After the transition week, the children started with an extended version of the FFD (FFD.E1), allowing lamb, butter and small portions of wheat, corn, potatoes, some fruits, and honey^7,18^. If no substantial behavioural improvement was reported, the FFD.E1 was adapted to FFD.E2 and FFD by gradually removing the additionally allowed foods. The most stringent FFD consisted of rice, turkey, vegetables (cabbage [white, green, Chinese, red], beet, cauliflower, borecole, swede, sprouts, lettuce), pears, olive oil, ghee, salt, rice drink with added calcium and water^19,20^. At t2, blood, urine, stool and buccal swap samples were collected and ADHD symptom score assessments were repeated.

### Behaviour scores

ADHD symptom scores were measured using the ADHD Rating Scale (ARS), consisting of 18 ADHD symptoms with a maximum score of 54^21,22^. The ARS was completed by the parents at t0, t1 and t2, focusing on the child’s behaviour during the past week. The child’s response to the FFD was determined with the percentage change in ARS score at t2 relative to t1 (100 × [t1 − t2] / t1). To estimate the response frequency to FFD, children with an ARS change ≥40% were designated responders, whereas children with <40% ARS change were designated non-responders^15^. Notably, in most statistical and molecular analyses reported in this manuscript the actual response percentages per child were used rather than the responder/non-responder classification.

### Sample collection and processing

Faecal samples were collected at t0, t1 and t2. Samples were collected in DNA/RNA Shield Faecal Collection Tubes (Zymo Research/BaseClear, Leiden, The Netherlands) and kept at room temperature until storage at −80 degrees Celsius. DNA was extracted according to the Standard Operating Procedure by Knudsen et al. 2016 (https://dx.doi.org/10.6084/m9.figshare.3475406), using the QIAmp Fast DNA Stool mini kit (Qiagen, Venlo, The Netherlands) and Lysing Matrix B beads (MP Biomedicals, Amsterdam, The Netherlands). DNA samples were frozen and shipped to Novogene (Cambridge, United Kingdom). For all samples, metataxonomic analyses were performed by V3-V4 16S rDNA amplicon sequencing using Illumina Novaseq, PE150 (30000 raw tags/sample). Metataxonomic 16S sequencing reads were processed with the R package DADA2 ^23^ and taxonomically assigned using the SILVA database v132^24^. Amplicon sequence variants (ASVs) with taxonomic assignments “eukaryote” and “chloroplast” were discarded. For the samples obtained at t1 and t2 (i.e., directly before and after the diet intervention period) shotgun metagenome datasets were generated using Illumina Novaseq, PE250 (40 milj. Reads/sample). Shotgun metagenome sequence reads were processed with HUManN2^25^. Relative bacterial species abundance and uniref90 gene families counts per million (CPM) were used for analyses. Unmapped reads (∼37% of total reads/sample) were removed before renormalization to CPM. In addition, for each sample reads were assembled into contigs with SPAdes version 3.14.1^26^ using the --meta option. Coverage was determined by mapping the reads back to these contigs as custom database in HUManN2. Abundance of the 21 gut microbial ECs involved in tyrosine and phenylalanine metabolism (i.e., 1.10.3.1, 1.14.16.1, 1.14.18.1, 1.3.1.43, 1.3.1.78, 1.3.1.79, 1.4.1.20, 1.4.3.2, 2.6.1.1, 2.6.1.5, 2.6.1.57, 2.6.1.58, 2.6.1.9, 4.1.1.25, 4.1.1.28, 4.1.99.2, 4.2.1.51, 4.2.1.91, 4.3.1.23, 4.3.1.24, 5.4.3.6) and of the KOs that make up the Raes modules^17^ were determined through HMM (Hidden Markov Model) search using HMMER 3.1b2 (http://hmmer.org/) against KOfam (HMM profiles for KEGG/KO with predefined score thresholds, https://www.genome.jp/ftp/db/kofam, downloaded on 26Aug2021). Abundances of Raes KOs stratified to gut microbial species were determined by regrouping the HUManN2 gene abundance table to KO abundance, keeping the taxa stratification.

Blood plasma and urine samples were collected at t1 and t2 and were available for 77 (t1) and 76 (t2) children. For 76 children both t1 and t2 samples were available. Blood samples were collected in BD Vacutainer® EDTA tubes (BD Biosciences, Vianen, The Netherlands), plasma was isolated by centrifugation 1300 x g, 10 minutes. Urine was collected and stored at −80 °C. Plasma and urine samples were shipped to Metabolon, Inc. (Morrisville, Unites States) for small-molecule profiling (HD4 Global Metabolomics platform).

Blood samples for PBMC isolation were collected at t1 and t2 and were available for 77 (t1) and 73 (t2) children. For 76 children both t1 and t2 samples were available. Blood samples for PBMC isolation were collected using BD Vacutainer® CPT™ Mononuclear Cell Preparation Tube - Sodium Citrate (BD Biosciences, Vianen, The Netherlands). PBMCs were isolated according to the manufacturer’s protocol, resuspended in TRIzol (Invitrogen) and stored at −80 °C. RNA was extracted according to the manufacturer’s protocol, resuspended in H_2_O and stored at −80 °C. RNA samples were shipped to Novogene (Cambridge, United Kingdom) for ribo-depleted RNA sequencing (Illumina Novaseq, PE150, 40 milj. raw reads/sample). Sequencing data was processed with the nf-core/rnaseq pipeline (<u>10.5281/zenodo.1400710</u>)^27^, applying the STAR-salmon option and mapping to the reference genome GRCh37.

Buccal swaps were collected at t1 and t2 using the Puritan™ HydraFlock™ Flocked Swabs (Merck Life Science NV, Amsterdam, The Netherlands). Collected samples were stored at −80 °C. Samples were transported to the HuGe-F facility of the Erasmus MC (Rotterdam, The Netherlands), where DNA methylation and SNP profiles were generated using the Infinium MethylationEPIC v1.0 (Legacy BeadChip, B5) and the Infinium™ Global Screening Array-24 v3.0 BeadChip, respectively. Infinium MethylationEPIC v1.0 data was processed using the R package meffil^28^.

### Data preprocessing

All datasets were reduced to have ∼5000 features or less, which required filtering of four data sets. 1) The faecal metagenomics genefamilies table was reduced from 2967553 to 4999 based on prevalence and CPM [Genefamilies with <42 CPM in <20 samples were removed]. 2) The PBMC RNA expression data was reduced from 57774 to 5003 genes based on prevalence and TPM (Transcripts Per Million) (length scaled) [Transcripts with <18 TPM in <13 samples were removed]. 3) The SNP data table was reduced from 654027 to 4785 features based on classification and prevalence [SNPs were removed from the table when they did not meet the two filtering criteria: i) being classified as nonsense or missense, and ii) prevalence of the non-dominate allele in ≥10 participants]. 4) The DNA methylation data table was reduced from 863560 to 5000 features based on the coefficient of variation (cv) for each feature, selecting the 5000 features with the highest cv (leading to the removal of all methylation features with a cv <0.62).

### Statistical analyses

Spearman rank correlation analysis was performed with the R package stats. Multi-omics factor analysis was performed using the MOFA2 package ^16,29^. MOFA is an unsupervised method for integrating multi-omics data producing latent factors across datasets. The standard data, model and training options were used for all models with the exceptions of not scaling the views and the convergence mode set to “slow”, and data were transformed with log(100*x+1). The number of learned factors was set to 10, accourding to the general MOFA guidelines (https://biofam.github.io/MOFA2/faq.html). Redundancy analysis was performed in Canoco version 5.12 ^30^, with default settings. In RDAs that included both t1 and t2 timepoints permutation was performed per participant to maintain coupling of the two measurements per individual. Differential gene expression analysis was conducted using the R package EdgeR ^31^, applying the default settings for the qCML approach to compare responders and non-responders. Differential gene expression data were analyzed using Ingenuity Pathway Analysis (IPA; QIAGEN, https://www.qiagenbioinformatics.com/products/ingenuity-pathway-analysis)^32^. IPA includes expertly curated biological interactions and functional annotations created from millions of individually modelled relationships between diverse molecular, cellular and clinical entities. The complete IPA output can be requested from the authors. As IPA input, HMDB metabolite abundance data and Ensembl ID gene expression data after EdgeR preprocessing to obtain fold-changes and p-values were provided and a Core Analysis with default settings was performed for urine (metabolomics) and blood plasma (metabolomics and gene expression) datasets. The output of IPA includes the overlap between the input data and (canonical) pathways, upstream regulators, diseases and cellular processes in Ingenuity’s Knowledge Base and returns two measures of association: (1) a ratio of the number of input metabolites that map to the pathway divided by the total number of metabolites that map to the same pathway, and (2) a p-value of the corresponding Fisher’s exact test to evaluate statistical support that the pathway might have been (significantly) modulated. From these and knowledge contained in the IPA Knowledge Base, IPA calculates z-scores that provide statistical support for determining if the modulated pathway was induced or repressed at the time of sample collection, and predicts molecules (cellular genes, proteins, regulatory RNAs, metabolites, pharmacological compounds) that might underpin the reported pathways and their corresponding z-scores. If the observed direction of change in a sufficiently large set of pathways or downstream genes (upstream regulators) is significantly consistent with a particular activation state of a pathway or transcriptional regulator, IPA predicts that the pathway or regulator had been “activated” or “inhibited”; such pathways and regulators have z-scores >2 or <-2. To facilitate comparing the most relevant IPA output, we used z-scores as cut-off and list the numbers of significantly activated or inhibited pathways and upstream regulators, with downstream target overlap at p-values <0.05.

## Data Availability

All data produced in the present study are available upon reasonable request to the authors

## Acknowledgements

This study was funded by a grant from Porticus. We thank all children and their parents for participating in this study. We thank the members of the Research Steering Committee, i.e., E. Aarts, M. Führer, T.R. Licht, D.J. Reijngoud, C.J.F. ter Braak, J. Toorman, M. de Boer, for advice and support.

## Author contributions

The authors’ responsibilities were as follows: M.K., R.R.P., and P.v.B. devised and designed the study. All authors contributed to data acquisition, analysis, and interpretation. S.H., J.B. and M.K. wrote the initial draft of the manuscript. All authors provided critical revision for intellectual content, approved the final version and were responsible for the decision to submit the manuscript.

