## Supplementary Material for "ADHD symptom reduction after following an FFD is associated with gut microbiome composition"

#### Supplementary Results

*Peripheral changes related to ARS change*

To investigate whether the change in ARS score between t1 and t2 (figure 2b) was related to changes in peripheral metabolites and gene expression, plasma metabolite profiles and gene expression levels of PBMCs at t2 corrected for baseline (t1) were investigated using the EdgeR software tool. Basically, the hypothesis that a beneficial decrease in ARS correlated significantly with differential expression of genes and/or abundance of specific metabolites was tested. When using multiple testing correction, EdgeR reported one differential metabolite (X – 25217) and no differentially expressed genes (DEGs) at pFDR<0.05. Since no significant relations were detected for individual genes and only one for the plasma metabolites, we extended the analysis to pathway enrichment, using the Ingenuity Pathway Analysis (IPA) software. IPA uses a knowledge base of more than 6 million validated interactions between genes, proteins and metabolites to calculate enrichment of pathways based on differential metabolites and gene expression data. When using as input, 770 differentially expressed genes and 25 metabolites (p(uncorrected)<0.05), IPA reported that 46 canonical pathways, involved in diverse cellular metabolic-, energy and immune-related pathways were significantly modulated (p<0.05), of which 2 were predicted (z-scores <-2 or >2; see Methods) to be repressed, i.e., representing pathways containing genes that correlated significantly with larger decreases in ARS. No pathways with z-scores >2 were reported. The 2 pathways with z-scores < 2 were "Assembly of RNA Polymerase II Complex" (p=0.021, z-score=-2.0) and "Oxidative Phosphorylation" (p=0.045, z=-2.4). IPA predicted that the observed differential gene expression patterns and the associated canonical pathways might be at least partially explained by activity of 8 activated regulators; repression of cellular pathways might have been controlled by inhibition of 10 regulators. Of these, regulators KDM5A, CEBPB and REL might have been involved in repression of pathway "Assembly or RNA polymerase II complex" These IPA results suggested that larger decreases in ARS score significantly correlated with differential (increased) expression of genes and concentrations of metabolites involved in gene expression and mitochondrial respiration.

### Supplementary Tables

**Table S1.** Association between ARS change (responders vs non-responders) and microbial genes of the phenylalanine and tyrosine synthesis pathways and phenylalanine and tyrosine levels in urine and plasma.

| | **Mean (sd)** | | | | **Association ARS change (parametric)[1]** | | | | **Association ARS (non-parametric)[2]** | |
|---|---|---|---|---|---|---|---|---|---|---|
| | **Responders** | | **Non-responders** | | | | | | | |
| **Microbial genes (EC)** | t1 | t2 | t1 | t2 | **p-value (t1)** | **p-value (ARS change[%])** | **FDR[3] (ARS change[%])** | **p-value (shapiro-Wilk.)** | **p-value** | **FDR[3]** |
| 1.3.1.43 | 2.31 (6.16) | 4.47 (6.25) | 1.97 (3.66) | 7.04 (11.14) | 5.03E-11 | 0.54 | 0.78 | 1.57E-05 | 0.73 | 0.95 |
| 2.6.1.1 | 283.21 (33.41) | 261.63 (35.25) | 280.67 (43.47) | 253.69 (35.89) | 0.04 | 0.38 | 0.62 | 0.01 | 0.94 | 0.98 |
| 2.6.1.57 | 0.7 (1.19) | 1.5 (2.71) | 1.63 (4.01) | 1.5 (2.41) | 1.78E-08 | 0.93 | 0.93 | 0.01 | 0.93 | 0.98 |
| 2.6.1.9 | 248.75 (33.37) | 238.87 (32.42) | 250.98 (42.8) | 221.83 (29.58) | 0.07 | 0.01 | 0.13 | 5.64E-03 | 0.11 | 0.48 |
| 4.1.1.25 | 0.05 (0.25) | 0.13 (0.42) | 0.12 (0.46) | 0.11 (0.48) | 4.65E-07 | 0.15 | 0.33 | 3.68E-03 | 0.03 | 0.20 |
| 4.1.1.28 | 0.12 (0.32) | 0.3 (0.55) | 0.01 (0.04) | 0.26 (0.57) | 1.98E-03 | 0.82 | 0.92 | 1.66E-13 | 0.41 | 0.78 |
| 4.1.99.2 | 0.28 (0.48) | 0.44 (0.66) | 0.54 (1.17) | 0.2 (0.41) | 0.34 | 0.07 | 0.30 | 1.15E-10 | 0.02 | 0.20 |
| 4.2.1.51 | 97.66 (31.42) | 114.49 (33.67) | 106.36 (39.62) | 119.56 (37.86) | 6.5E-04 | 0.84 | 0.92 | 0.02 | 0.59 | 0.95 |
| 4.2.1.91 | 2.86 (2.22) | 3.63 (3.39) | 3.19 (2.38) | 3.78 (2.23) | 1.69E-12 | 0.85 | 0.92 | 0.82 | 0.98 | 0.98 |
| **Metabolites** | | | | | | | | | | |
| Phenylalanine | | | | | | | | | | |
| *Plasma* | 1.01 (0.12) | 1.00 (0.12) | 0.96 (0.09) | 1.05 (0.12) | 0.54 | 0.14 | 0.33 | 0.02 | 0.25 | 0.78 |
| *Urine* | 1.03 (0.28) | 1.13 (0.3) | 1.00 (0.24) | 1.02 (0.37) | 0.8 | 0.1 | 0.33 | 0.73 | 0.40 | 0.78 |
| Tyrosine | | | | | | | | | | |
| *Plasma* | 1.08 (0.18) | 0.94 (0.16) | 1.03 (0.2) | 1.01 (0.22) | 0.63 | 0.18 | 0.33 | 0.03 | 0.73 | 0.95 |
| *Urine* | 1.07 (0.36) | 1.08 (0.36) | 1.06 (0.36) | 0.93 (0.37) | 0.39 | 0.04 | 0.26 | 0.32 | 0.42 | 0.78 |

[1] ANCOVA: t2 ~ t1 + ARS change (responder vs non-responder)
[2] Mann-Whitney test: responders log2(t2/t1) vs non-responders log2(t2/t1)
[3] Benjamini & Hochberg

**Table S2.** Association between ARS change (%) and microbial genes of the phenylalanine and tyrosine synthesis pathways and phenylalanine and tyrosine levels in urine and plasma.

| | | | **Association ARS change (parametric)[1]** | | | | **Association ARS (non-parametric)[2]** | |
|---|---|---|---|---|---|---|---|---|
| | **t1 Mean (sd)** | **t2 Mean (sd)** | **p-value (t1)** | **p-value (ARS change[%])** | **FDR[3] (ARS change[%])** | **p-value (shapiro-Wilk.)** | **p-value** | **FDR[3]** |
| **Microbial genes (EC)** | | | | | | | | |
| 1.3.1.43 | 2.19 (5.35) | 5.41 (8.41) | 0.01 | 0.13 | 0.28 | 3.67E-10 | 0.62 | 0.90 |
| 2.6.1.1 | 282.28 (37.17) | 258.71 (35.46) | 0.16 | 0.12 | 0.28 | 0.23 | 0.96 | 0.96 |
| 2.6.1.57 | 1.04 (2.62) | 1.5 (2.59) | 0.05 | 0.64 | 0.80 | 3.20E-11 | 0.91 | 0.96 |
| 2.6.1.9 | 249.57 (36.85) | 232.61 (32.29) | 0.12 | 0.01 | 0.13 | 9.32E-06 | 0.18 | 0.59 |
| 4.1.1.25 | 0.08 (0.34) | 0.13 (0.44) | 2.98E-06 | 0.74 | 0.80 | 3.84E-16 | 0.13 | 0.56 |
| 4.1.1.28 | 0.08 (0.26) | 0.28 (0.56) | 0.01 | 0.94 | 0.94 | 2.03E-13 | 0.62 | 0.90 |
| 4.1.99.2 | 0.37 (0.81) | 0.36 (0.59) | 0.76 | 0.06 | 0.26 | 5.68E-09 | 0.02 | 0.26 |
| 4.2.1.51 | 100.85 (34.66) | 116.35 (35.11) | 1.75E-03 | 0.69 | 0.80 | 0.34 | 0.32 | 0.83 |
| 4.2.1.91 | 2.98 (2.27) | 3.68 (3.01) | 7.80E-07 | 0.50 | 0.72 | 3.17E-08 | 0.47 | 0.87 |
| **Metabolites** | | | | | | | | |
| Phenylalanine | | | | | | | | |
| *Plasma* | 0.99 (0.11) | 1.02 (0.12) | 0.53 | 0.21 | 0.36 | 0.01 | 0.08 | 0.52 |
| *Urine* | 1.02 (0.26) | 1.09 (0.33) | 0.76 | 0.22 | 0.36 | 0.47 | 0.39 | 0.85 |
| Tyrosine | | | | | | | | |
| *Plasma* | 1.06 (0.19) | 0.96 (0.19) | 0.65 | 0.08 | 0.26 | 0.03 | 0.83 | 0.96 |
| *Urine* | 1.07 (0.36) | 1.02 (0.37) | 0.44 | 0.049 | 0.26 | 0.41 | 0.80 | 0.96 |

[1] ANCOVA: t2 ~ t1 + ARS change (%)
[2] Spearman rank: microbial genes (EC) or metabolites log2(t2/t1) vs ARS change (%)
[3] Benjamini & Hochberg

#### Supplementary Figures

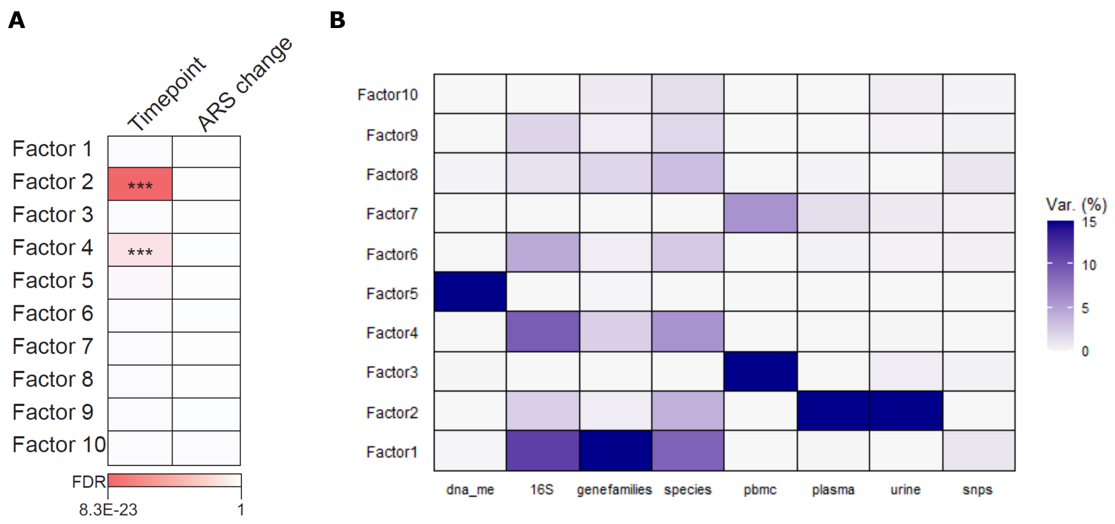


**Figure S1.** 10-factor MOFA model, FFD only (n=68). A) Associations of the 10 factors in the MOFA model with timepoint (t1 vs t2) and ARS change (%). B) Explained variation (%) of the data views per factor.

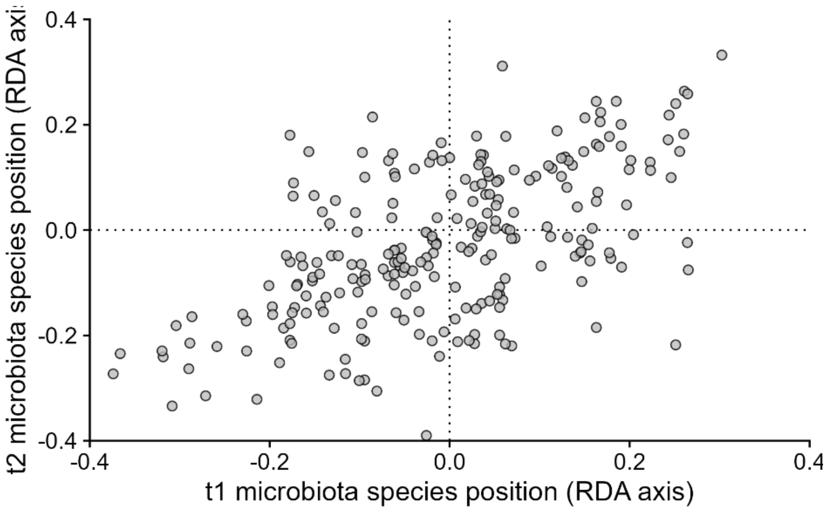


**Figure S2.** Scatterplot of the t1 and t2 microbiome species positions on the ordination axes of the RDAs with ARS change as explanatory variable and gut microbiome species composition at t1 ($p=0.042$, explained variation=2.13%) or t2 ($p=0.006$, explained variation=2.40%) as response variables (FFD only, n=68).

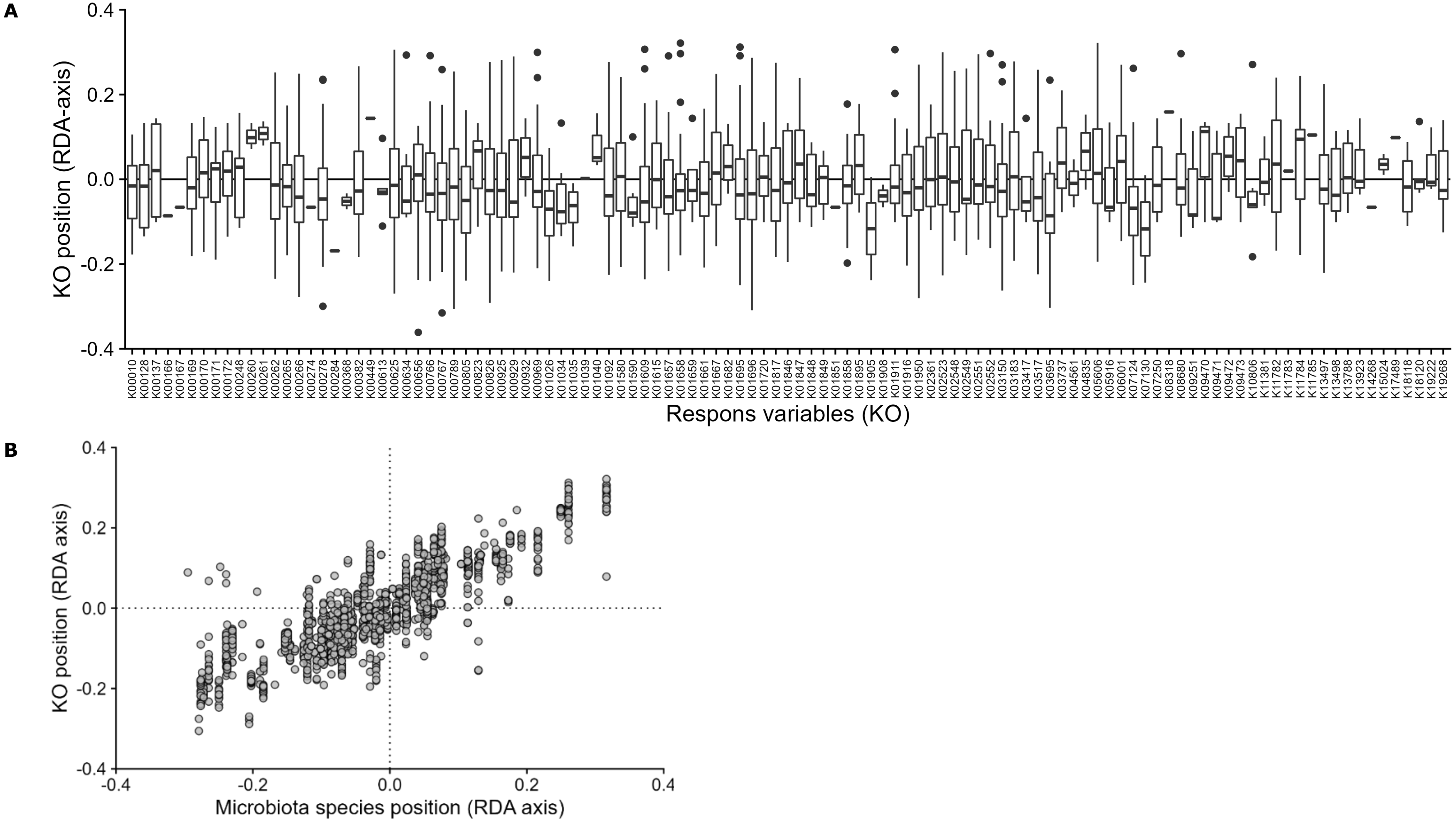


**Figure S3.** Positions of the KO response variables in the RDA ordination axis of the RDA with ARS change (%) as explanatory variable and relative KO abundance stratified to species (figure 5b) or relative species abundance (figure 5a) as response variables. A) Boxplot of KO positions per KO (x-axis) stratified to microbiota species by which they are encoded (y-axis). B) Scatterplot of KO positions stratified to microbiota species (y-axis) and species microbiota positions of the RDA of the RDA with relative species abundance as response variable.

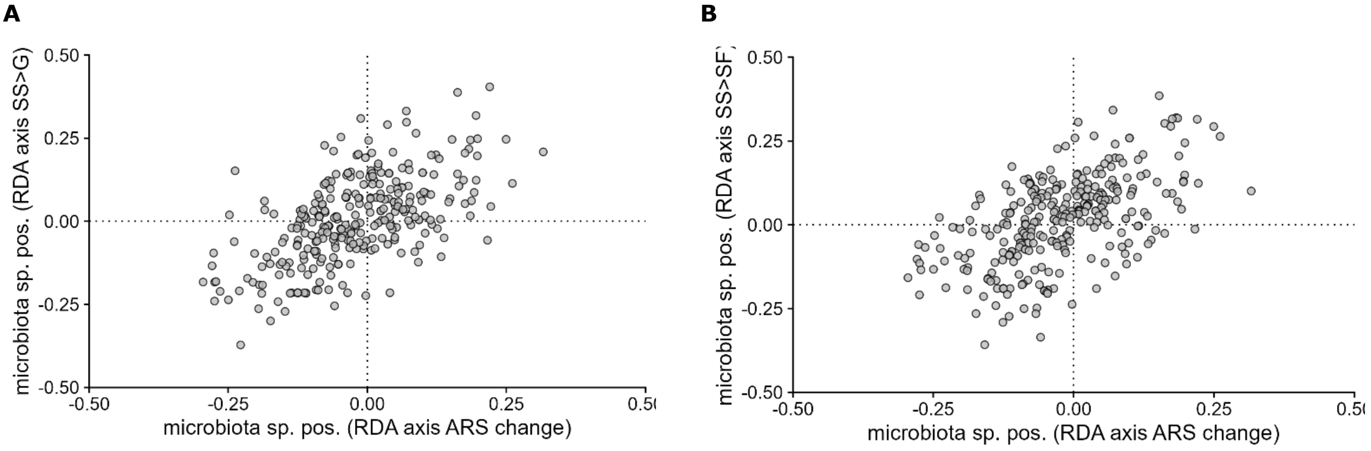


**Figure S4**. Scatterplots of the microbiota species positions on the RDA axis in the RDAs with ARS change as explanatory variable versus t2-t1 beta weights of the A) StopSuccess>Go or B) StopSuccess>StopFail contrasts.

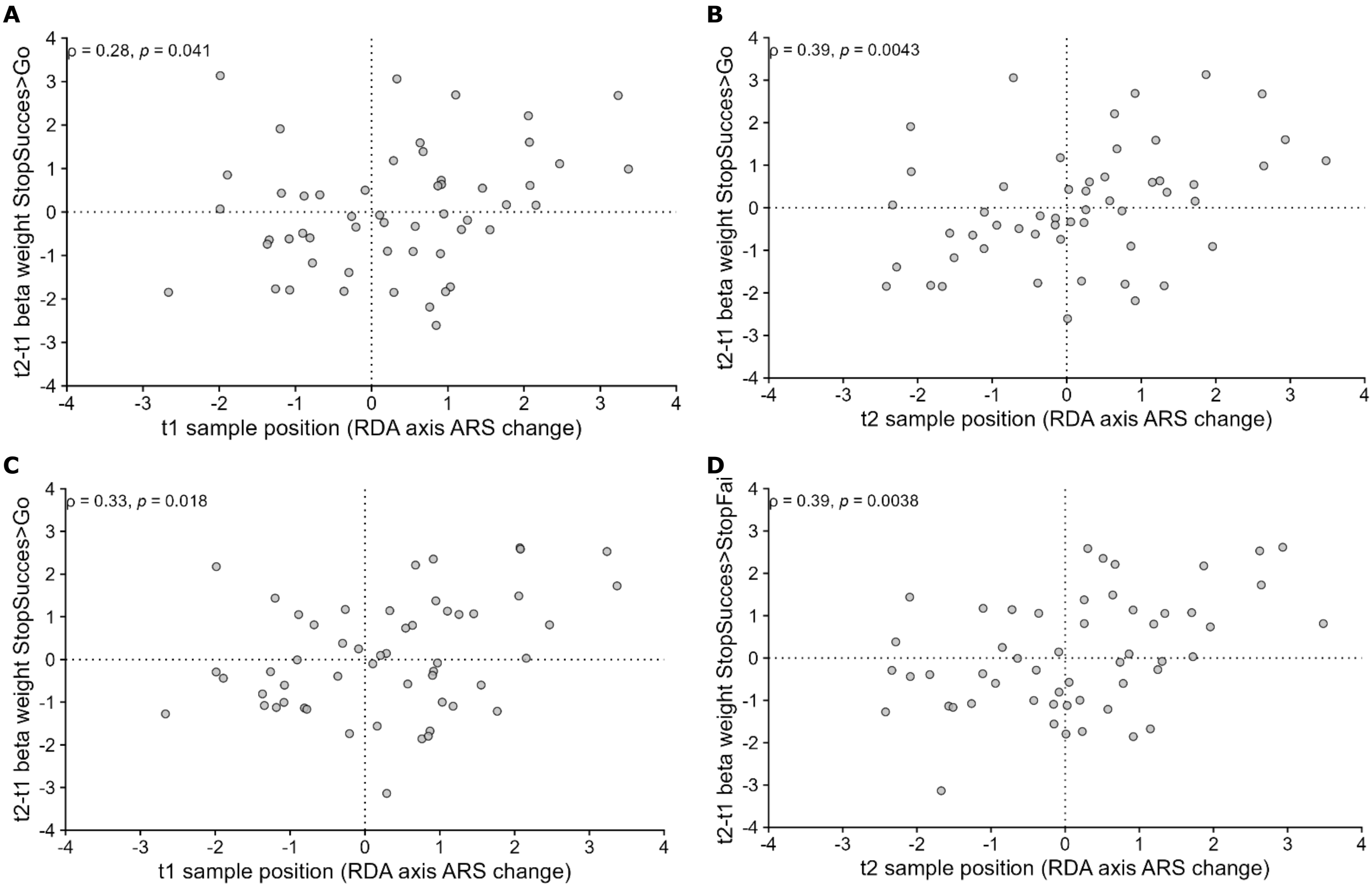


**Figure S5.** Scatterplots and Spearman rank p-values of the precuneus t2-t1 beta weights of A-B) the StopSuccess>Go or C-D) the StopSuccess>StopFail contrasts versus the RDA axis position of A,C) the t1 samples or B,D) the t2 samples from the RDA with microbiota species abundance as response variable and ARS change as explanatory variable (figure 5a).
